# The Effect of Neutropenia and Filgrastim (G-CSF) in Cancer Patients With COVID-19 Infection

**DOI:** 10.1101/2020.08.13.20174565

**Authors:** Sejal Morjaria, Allen W Zhang, Anna Kaltsas, Rekha Parameswaran, Dhruvkumar Patel, Wei Zhou, Jacqueline Predmore, Rocio Perez-Johnston, Justin Jee, Miguel-Angel Perales, Anthony F. Daniyan, Ying Taur, Sham Mailankody

## Abstract

**Background:** Neutropenia is commonly encountered in cancer patients, and recombinant human granulocyte colony-stimulating factor (G-CSF, filgrastim) is widely given to oncology patients to counteract neutropenia and prevent infection. G-CSF is both a growth factor and cytokine that initiates proliferation and differentiation of mature granulocytes. However, the clinical impact of neutropenia and G-CSF use in cancer patients, who are also afflicted with coronavirus disease 2019 (COVID-19), remains unknown.

**Methods:** An observational cohort of 304 hospitalized patients with COVID-19 at Memorial Sloan Kettering Cancer Center was assembled to investigate links between concurrent neutropenia (N=55) and G-CSF administration (N=16) on COVID-19-associated respiratory failure and death. These factors were assessed as time-dependent predictors using an extended Cox model, controlling for age and underlying cancer diagnosis. To determine whether the degree of granulocyte response to G-CSF affected outcomes, a similar model was constructed with patients that received G-CSF, categorized into “high”- and “low”- response, based on the level of absolute neutrophil count (ANC) rise 24 hours after growth factor administration.

**Results:** Neutropenia (ANC < 1 K/mcL) during COVID-19 course was not independently associated with severe respiratory failure or death (HR: 0.71, 95% Cl: 0.34-1.50, *P* value: 0.367) in hospitalized COVID-19 patients. When controlling for neutropenia, G-CSF administration was associated with increased need for high oxygen supplementation and death (HR: 2.97, 95% CI: 1.06-8.28, *P* value: 0.038). This effect was predominantly seen in patients that exhibited a “high” response to G-CSF based on their ANC increase post-G-CSF administration (HR: 5.18, 95% CI: 1.61-16.64, *P* value: 0.006).

**Conclusion:** Possible risks versus benefits of G-CSF administration should be weighed in neutropenic cancer patients with COVID-19 infection, as G-CSF may lead to worsening clinical and respiratory status in this setting.

## Introduction

Neutropenia is a common side-effect of anti-cancer therapies, and recombinant human granulocyte colony stimulating factor (G-CSF, filgrastim) is often given to cancer patients for ongoing or impending neutropenia. During the ongoing SARS-CoV-2 pandemic, there is uncertainty about the effect of commonly used medications like G-CSF on clinical outcomes. The mechanism of action of G-CSF (stimulates both cytokine and neutrophil production),^1^ the association of G-CSF with acute lung injury (ALI) or adult respiratory distress syndrome (ARDS),^2,3^ and what is now known about the pathogenesis of COVID-19- resulting in cytokine storm in some patients, has raised concerns about the safety profile of G-CSF in COVID-19 patients. Moreover, lung findings from autopsies of COVID-19 patients have shown neutrophil extravasation in the alveolar space,^4,5,6^ raising concerns that G-CSF administration and the resultant neutrophil expansion, could to lead to exaggerated neutrophil responses, thereby worsening clinical outcomes in COVID-19 patients.

Our group published a case series describing the rapid clinical deterioration of three COVID-19 patients soon after receiving G-CSF. Herein, we sought to quantitatively estimate the clinical effects of G-CSF on a larger cohort of COVID-19 patients.^7^ Specifically, we asked: what are the effects of neutropenia and G-CSF administration on the clinical outcomes, respiratory failure or death, in patients with cancer and COVID-19 infection.

## Methods

### Study population

We included all 304 inpatients admitted within a window of [-5 days, +14 days] around a patient’s COVID-19-postive test date (day 0), between the dates of March 13, 2020 and May 15, 2020. Clinical outcomes were monitored until May 19, 2020. If a patient reached a clinical endpoint (defined below) prior to receiving G-CSF, they were excluded from the cohort. Patients with a diagnosis of acute myeloid leukemia or myelodysplastic syndrome were not included in the analysis, as these patients are generally not candidates for G-CSF administration. As we were primarily interested in whether or not G-CSF should be administered for neutropenia in the context of a COVID-19 infection, patients that received G-CSF before the first recorded occurrence of neutropenia (defined as ANC < 1 K/mcL) or prior to hospitalization were excluded from the analysis. Furthermore, when stratifying patients that received G-CSF into high and low responders, patients without ANC values on day 0 and day 1 of G-CSF administration were also excluded from the analysis, as a log-fold change in ANC could not be computed for these patients. We define G-CSF administration as the inpatient use of filgastrim or pegfilgastrim at any dose. The Memorial Sloan Kettering Cancer Center (MSKCC) institutional review board approved the study.

### Laboratory methods

COVID-19 status was determined using a nasopharyngeal swab to determine the presence of virus specific RNA (MSKCC FDA EUA-approved assay and Cepheid^®^). COVID-19 RNA was detected using the Centers for Disease Control and Prevention protocol targeting two regions of the nucleocapsid gene (N1 and N2) with modifications described elsewhere. ^8-10^

### Data sources

Patient data was extracted from the MSKCC electronic health record. Patient medications, demographics, and outcomes (i.e. use of high-flow supplemental oxygen, mechanical ventilation or tracheostomy, and death), were extracted from a standardized-input institutional database. Respiratory failure was defined as oxygen supplementation of ≥ 4 liters per minute on nasal cannula, high-flow nasal cannula, or any amount of oxygen on non-rebreather, Bi-Level Positive Airway Pressure (BIPAP), or mechanical ventilation.

### Statistical analysis

In the primary analysis, we applied an extended Cox model (Cox regression model) using age (binned into < 25, 25-50, 50-70, 70-80, 80-90, and >90 categories) and cancer type as time-independent covariates. The first occurrence of neutropenia and the first administration of G-CSF (filgrastim or pegfilgrastim) following COVID-19 positivity were introduced as binary time-dependent covariates in the model. To minimize reverse causality, and since the earliest evidence of ANC recovery we had was a day after G-CSF administration with few changes in the successive days, G-CSF events were encoded 1 day following actual administration, to capture the timing of its expected effect. In a second analysis, G-CSF response was additionally categorized into “high-response” and “low-response” groups and introduced as mutually exclusive binary time-dependent covariates. High response was defined as a > 50th percentile increase in ANC at day 1 post-G-CSF administration (compared to ANC values prior to G-CSF administration) across all cases of G-CSF administration. For our cohort, this corresponded to an increase in ANC of at least fourfold from day 0 to day 1 after G-CSF administration.

For survival analysis, starting time was defined as the time of COVID-19 diagnosis for each patient. To verify that the significant hazard ratio associated with G-CSF administration was not solely due to collinearity between the neutropenia and G-CSF variables, we also considered an alternative formulation of the model, where starting time was instead defined as the time of first occurrence of neutropenia. The number of days prior to neutropenia was added as a continuous time-independent variable to the model. The primary analysis was conducted using a composite endpoint defined as the first occurrence of respiratory failure (defined above) or death following COVID-19 diagnosis. For the alternate analysis, we excluded death as an endpoint and only looked at patients that developed respiratory failure, as patients that died without oxygen supplementation (n = 3) may have died of non-COVID-19-related causes.

All patient events were right-censored at the date of last follow-up or May 19, 2020. All analysis was performed in R version 3.6.2 with the survival (version 3.1-11) and survminer (version 0.4.7) packages. ^11^

## Results

We assembled a cohort of 304 hospitalized patients with cancer who tested positive for COVID-19 at MSKCC between the dates of March 13, 2020 and May 15, 2020. These cases comprised a variety of cancer types, including gastrointestinal malignancies (n=45, 14.8%), lung cancers (n=41, 13.5%), non-Hodgkin lymphomas (n=39, 12.8%), and breast cancers (n=37, 12.2%), among others. In total, 55 (18.1%) patients were neutropenic at some point during hospitalization, as defined by an ANC less than 1 K/mcL. Of these, 16 (48.4%) of patients received either filgrastim or pegfilgrastim (G-CSF) for neutropenia. A total of 103 (33.9%%) patients reached the primary endpoint: the development of respiratory failure. The patient characteristics of all patients (n=304) are shown in **Table 1**.

**Table 1.** Demographics and baseline characteristics of all inpatients with COVID-19 (n = 304)

| **Characteristic** | **Number of Patients, n (%)** |
| --- | --- |
| **Age (years)** |  |
| < 18 years | 10 (3.29) |
| 18-29 | 3 (0.99) |
| 30-39 | 6 (1.97) |
| 40-49 | 34 (11.2) |
| 50-59 | 59 (19.4) |
| 60-69 | 90 (29.6) |
| 70-80 | 70 (23.03) |
| 80-90 | 27 (8.88) |
| > 90 | 5 (1.64) |
| **Gender** |  |
| Male | 144 (47.4) |
| Female | 160 (52.6) |
| **Race** |  |
| White | 182 (59.9) |
| Black | 55 (18.1) |
| Asian | 23 (7.57) |
| Other | 44 (14.5) |
| **Underlying Cancer** |  |
| Breast cancer | 37 (12.2) |
| CNS cancers<sup>+</sup> | 6 (1.97) |
| Germ cell tumors<sup>Ⓛ</sup> | 3 (1.0) |
| Lung cancer | 41 (13.5) |
| Gastrointestinal malignancies<sup>∞</sup> | 45 (14.8) |
| Genitourinary malignancies<sup>Δ</sup> | 33 (10.9) |
| Gynecologic malignancies<sup>√</sup> | 15 (4.93) |
| Head and neck cancers<sup>Θ</sup> | 7 (2.3) |
| Melanoma | 4 (1.32) |
| Sarcoma | 11 (2.1) |
| Acute and chronic leukemias<sup>†</sup> | 2 (6.2) |
| Plasma cell dyscrasias<sup>‡</sup> | 25 (8.6) |
| Myeloproliferative neoplasms<sup>▷</sup> | 7 (2.3) |
| Non-Hodgkin lymphoma | 39 (12.8) |
| Hodgkin lymphoma | 5 (1.64) |
| Other | 5 (1.64) |
| **Neutropenia (ANC ≤ 1)** | **55 (18.1)** |
| **All inpatient G-CSF use** |  |
| Filgrastim | 25 (8.22) |
| Pegylated filgrastim | 4 (1.32) |
| Neither | 275 (90.5) |
| **Inpatient G-CSF use prior to combined endpoint only** |  |
| Filgrastim | 15 (4.93) |
| Pegylated filgrastim | 1 (0.3) |
| Neither | 288 (94.7) |
| **Supplemental O2\*\*** |  |
| Room air | 105 (34.5) |
| Low O2 (≤ 4 L of O2) | 45 (14.8) |
| High O2 (> 4 L of O2)*** | 103 (35.3) |
| Mechanical Ventilation | 51 (16.8) |
| **Death** | **53 (17.4)** |

\*\* Highest oxygen requirement during hospitalization
\*\*\* Includes high flow nasal oxygen, non rebreather mask, bilevel positive airway pressure
**CNS cancers<sup>+</sup>** = Astrocytoma, glioblastoma, glioma, nerve sheath tumor, neuroblastoma
**Germ cell tumors<sup>Ⓛ</sup>** = Germ cell tumor and testicular cancer
**Gastrointestinal malignancies<sup>∞</sup>** = Appendiceal, cholangiocarcinoma, gallbladder, gastrim, esophageal, hepatocellular, pancreatic, rectal/colon
**Genitourinary malignancies<sup>Δ</sup>** = Bladder cancer, urothelial cancer, prostate cancer, renal cell carcinoma
**Gynecologic malignancies<sup>√</sup>** = cervical, endometrial, ovarian, uterine, vulvar
**Head and neck cancers<sup>Θ</sup>** = squamous cell cancer (head and neck), adenoid cystic carcinoma, thyroid
**Acute and chronic leukemias<sup>†</sup>** = acute lymphoblastic leukemia, acute myeloid leukemia, chronic lymphoid leukemia, chronic myeloid leukemia, chronic myelomonocytic leukemia, hairy cell leukemia, myelodysplastic syndrome
**Plasma cell dyscrasias<sup>‡</sup>** = amyloidosis, multiple myeloma, monoclonal gammopathy of undetermined significance
**Myeloproliferative neoplasms<sup>▷</sup>** = aplastic anemia, immune thrombocytopenia purpura, Erdheim Chester, myelofibrosis.

The key clinical timepoints for patients (neutropenia, G-CSF administration, respiratory failure, or death) for each patient (*prior to applying exclusion criteria) above are shown in **Figure 1**. The patient characteristics of neutropenic inpatients that did (n=29) and did not receive G-CSF (n=55) are shown in **Table S1**. Patient characteristics of both inpatients and outpatients with COVID-19 infection that did and did not receive G-CSF with their clinical outcomes are shown in **Tables S2-S3**. A total of nine of 29 inpatients (31%) receiving G-CSF before application of exclusion criteria required >4L oxygen, compared with 94 of 275 (34%) patients not receiving G-CSF.

**Figure 1:**
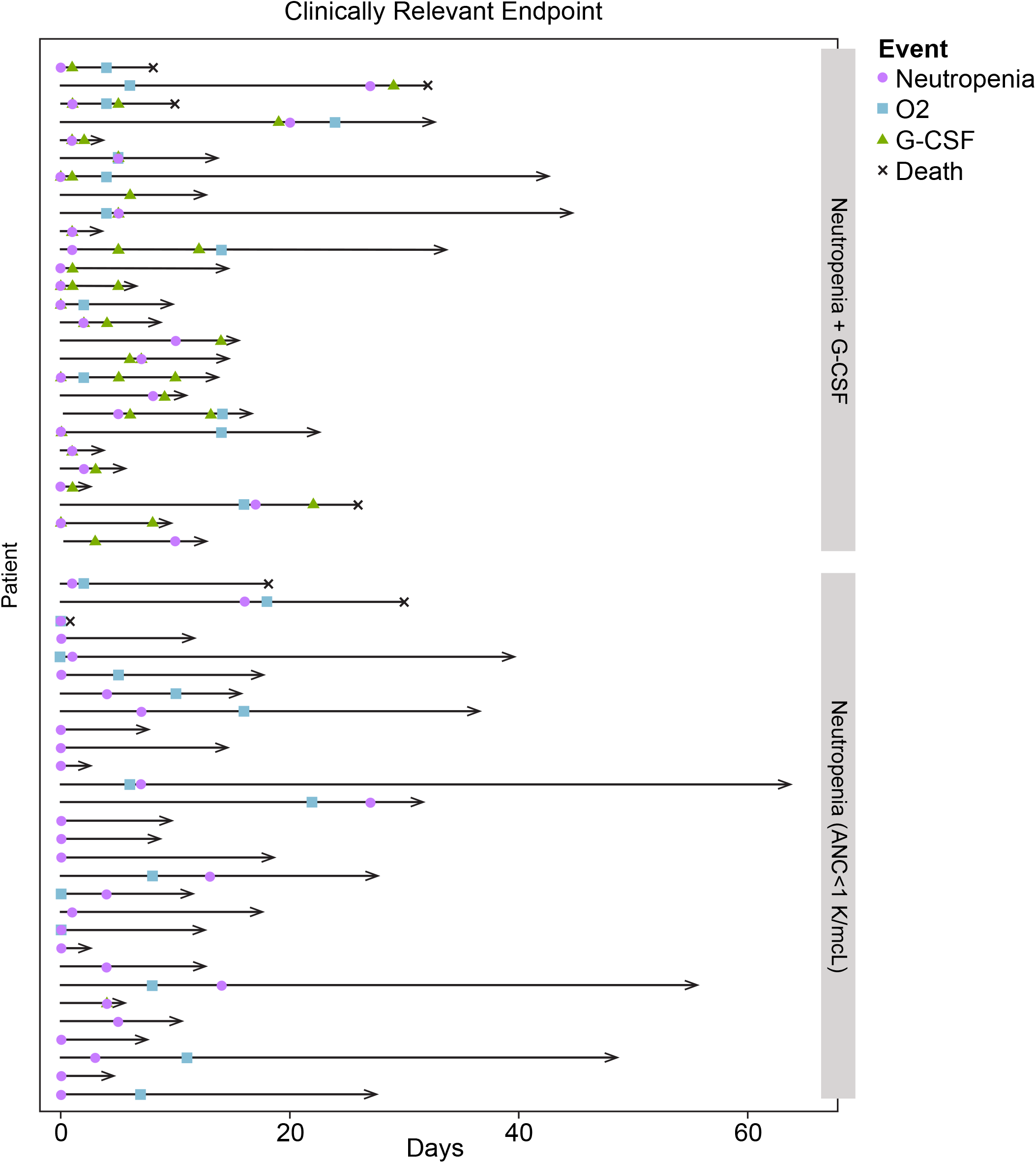
Timelines of all hospitalized, neutropenic (ANC < 1 K/mcL) patients who received Granulocyte-colony-stimulating factor (G-CSF (top panel)) versus those patients that were neutropenic but did not receive G-CSF (bottom panel) showcasing their relevant clinical endpoints (neutropenia, G-CSF administration, “respiratory failure” (defined in Methods). Three patients that received G-CSF as an outpatient, subsequently admitted to the hospital were counted in subsequent analysis as being neutropenic but not receiving G-CSF.

A total of 16 patients were counted as having received G-CSF, based on our definition. To evaluate the effect of neutropenia and G-CSF use on our primary composite endpoint defined as respiratory failure and death from COVID-19, we applied an extended Cox model controlling for age and neutropenia while stratifying by underlying cancer diagnosis (see Methods). As expected, age at enrollment greater than 90 (HR: 7.29, 95% CI: 2.72-19.52, *P* value: <0.001) was associated with significantly worse outcomes. In contrast, being less than 50 years of age was associated with better outcomes. G-CSF use (HR: 2.97, 95% CI: 1.06-8.28, *P* value: 0.038; **Figure 2**), but not neutropenia alone (HR: 0.71, 95% CI: 0.34-1.49, p value: 0.364), portended significantly worse outcomes in a multivariate model. These results were consistent with those obtained using respiratory failure alone as the endpoint (HR: 3.01, 95% CI: 1.08-8.39, *P* value: 0.035; **Figure S1**), or when considering only neutropenic patients and using time of neutropenia as t=0 (HR for G-CSF: 4.62, 95% CI: 1.08-19.7, *P* value: 0.039; **Figure S2**).

**Figure 2:**
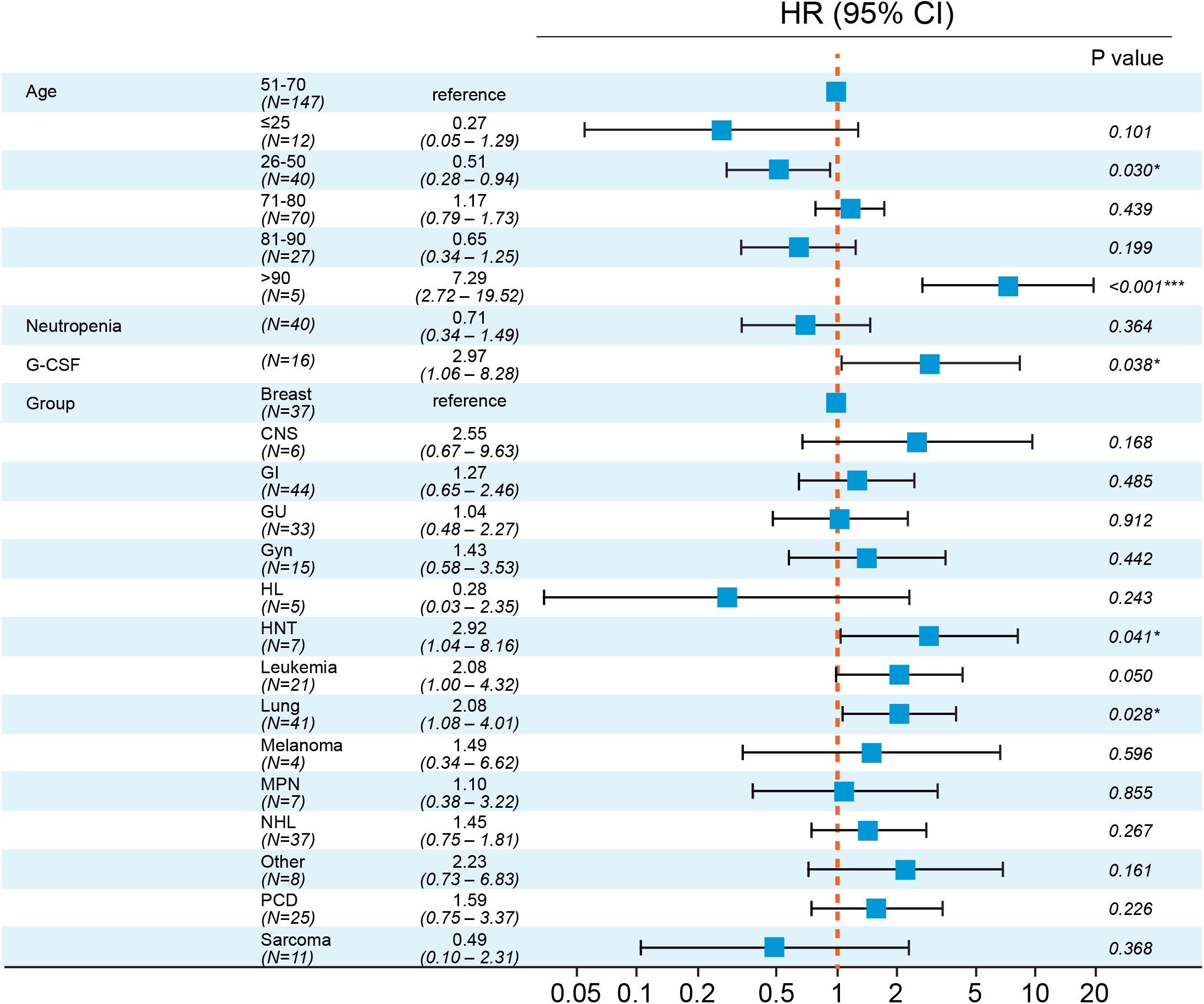
Forest plot showing the effect (HR = Hazard Ratio) of Granulocyte colony-stimulating factor (G-CSF) on the composite endpoint of the first occurrence of “respiratory failure” (defined in Methods) or death. Hazard ratios were computed with an extended Cox model, using binned ages and cancer type as time-independent covariates and neutropenia and G-CSF as time-dependent covariates.

To determine whether the neutrophil-inducing properties of G-CSF may relate to the poor outcomes associated with G-CSF, we further considered the neutrophil concentrations in peripheral blood prior to and immediately after G-CSF administration. As expected, ANC and ANC/ALC (ratio of absolute neutrophil count to absolute lymphocyte count) values increased after G-CSF administration, and this increase was predominantly limited to the day immediately following G-CSF administration (**Figure 3A,3C**). In contrast, ALC values remained more constant during this 1-day period (**Figure 3B**), and thus the rise in ANC/ALC is largely attributable to ANC. We stratified patients that received G-CSF (n=16) based on their response to ANC, computed as the log fold-change between ANC values 1 day after versus the day of G-CSF (day 0) administration (**Figure 4A**). High-response patients (n=9; **Figure 4B**) were defined as those that experienced a >4X rise in ANC, comprising the upper 50th percentile of the patients that received G-CSF. Using these response categories, we modified the extended Cox model shown in Figure 2 to categorize G-CSF events as high- and low- response. As before, age greater than 90 (HR: 7.37, 95% CI: 2.75-19.76, *P* value<0.001) was significantly associated with worse outcomes.

**Figure 3.**
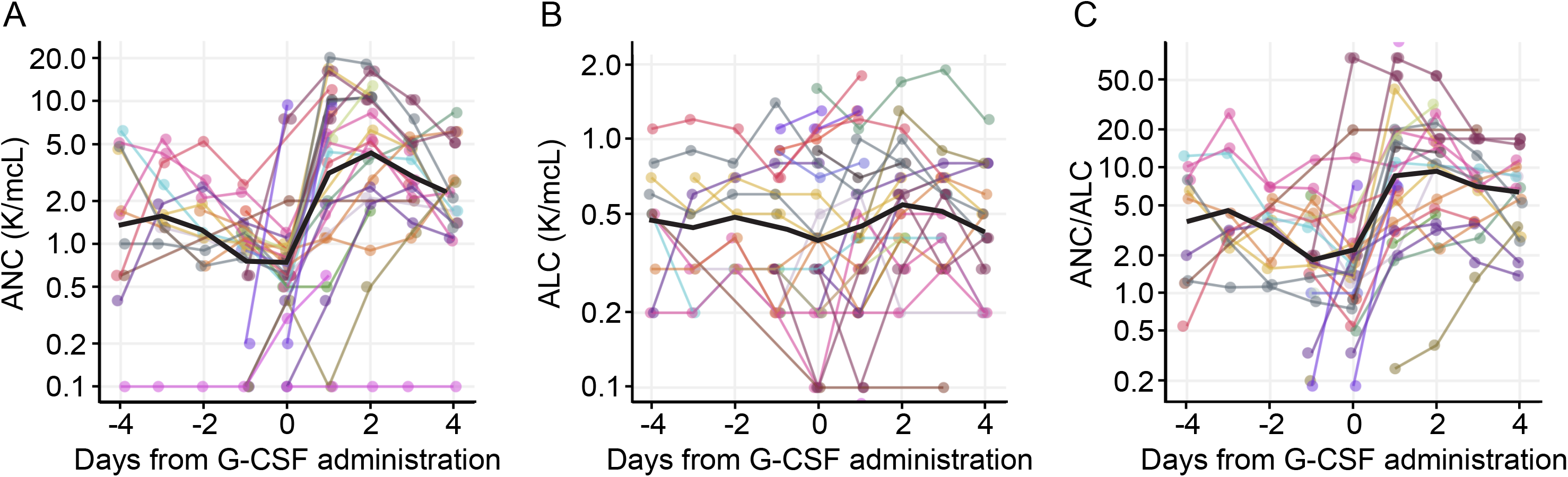
Lab values of: Absolute Neutrophil Count (ANC (K/mcL)) **(a)**, Absolute Lymphocyte Count (ALC (K/mcL)) **(b)**, and their ratio, ANC/ALC **(c)** within a 4 day window of G-CSF administration. Day 0 corresponds to the date of G-CSF administration. The black dashed line in each panel corresponds to the average lab value per day for all patients.

**Figure 4:**
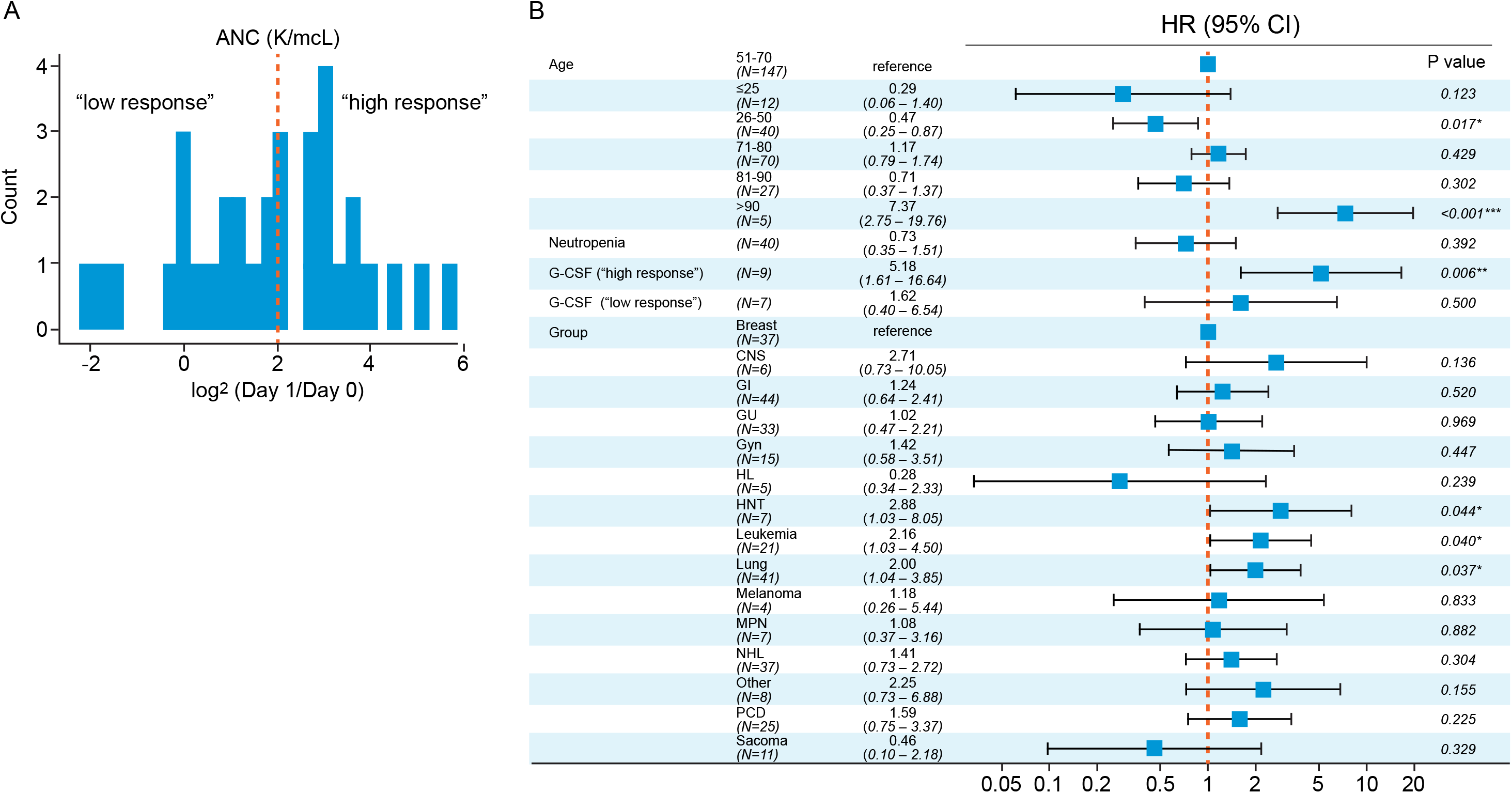
**(a)** Log-fold change values of absolute neutrophil count (ANC) obtained 1 day after G-CSF administration and prior to G-CSF administration for the n=25 patients that received G-CSF. Patients were stratified into “high” (to the right of the dashed orange line; fourfold increase in ANC day 1 post-G-CSF) or “low” (to the left of the dashed red line) responders based on these values (threshold of ANC = 2, corresponding to the 50th percentile). **(b)** Forest plot showing the effect (HR = Hazard Ratio) of high- and low-response to G-CSF on the first occurrence of “respiratory failure” (defined in Methods). Hazard ratios were computed with an extended Cox model, using binned ages and cancer type as time-independent covariates, and neutropenia and G-CSF (high- and low-response) as time-dependent covariates.

There were pronounced differences in risk associated with low- and high-response to G-CSF. While low response to G-CSF was not significantly associated with poorer outcomes (HR: 1.62, 95% CI: 0.40 - 6.54, *P* value: 0.5), high response to G-CSF was significantly associated with poorer outcomes (HR: 5.18, 95% CI: 1.61-16.64, *P* value: 0.006; **Figure 4B**). These results imply that the effect of G-CSF on outcomes is primarily driven by patients with robust increases in ANC following G-CSF administration. Interestingly, neutropenia was not statistically associated with worse outcomes.

Ten of the 16 patients that were neutropenic and received G-CSF had a pre-and-post chest radiograph and 6/10 (60%) demonstrated radiologic worsening within 7 days of receiving G-CSF (**Table S4**); 3 patients that developed radiographic deterioration post-G-CSF use are shown in **Figure S3**.

## Discussion

The COVID-19 pandemic has made delivering effective cancer care -- already a challenging endeavor -more difficult, given the balancing of competing risks of death from untreated cancer versus death or serious complications from COVID-19 infection.^12, 13, 14, 8 15, 16^ In this study, we evaluate the potential impact of neutropenia and G-CSF use in cancer patients with concurrent COVID-19 infection. We observed a higher likelihood of respiratory failure and death in patients that received G-CSF, particularly in the subset of patients that exhibited a “high” response to G-CSF. Our observations also suggest that neutropenia during COVID-19 illness itself was not an independent risk factor for adverse outcomes in COVID-19 illness.

From published reports, it is becoming increasingly clear that rapid clinical deterioration can occur in some patients because of the hyperactive immune response driving COVID-19 progression, causing an overwhelming infiltration of inflammatory myeloid cells into the lungs (particularly monocytes, macrophages and neutrophils),^4,17–19^ the so-called “cytokine storm”. Zuo et al. used cell-free DNA as a marker to detect neutrophil extracellular trap (NET) remnants in the blood and noted that these appeared strongly correlated with absolute neutrophil counts, and elevated blood neutrophils forecast worst outcomes. ^6^ Lung injury is one consequence of the cytokine storm that can progress into ALI or its more severe form, ARDS.^20^ Considering that neutrophil influx in the lung is a hallmark feature of ARDS,^21^ and that ALI has already been reported as a potential complication of G-CSF use, ^2,3^ administering G-CSF to certain cancer patients with COVID-19 for neutropenia may give clinicians pause. Similar concerns exist for patients who receive chimeric antigen receptor T (CAR-T) cell therapy,^22^ where G-CSF administration is generally avoided to prevent the overactivation of the immune system, given the already increased risk for cytokine storm.

To best estimate the effects of neutropenia and G-CSF on our defined clinical outcomes, we encoded G-CSF as a time-dependent covariate, while controlling for neutropenia as another time-dependent covariate. We also compared the clinical outcomes of patients that had different levels of response to G-CSF, finding that robust G-CSF neutrophil response was associated with substantially higher hazard (HR: 5.18, 95% CI: 1.61-16.64, *P* value: 0.006) for respiratory decompensation, compared to those that had less robust levels of response to G-CSF (HR: 1.62, 95% CI: 0.40-6.54, *P* value: 0.500). We also show that soon after G-CSF administration, there is a substantial increase in the neutrophil to lymphocyte ratio (ANC/ALC), previously shown to be an independent risk factor for mortality in hospitalized patients with COVID-19. ^23^ To our knowledge, this is the first study describing the course of COVID-19 infection in selected cancer patients who received G-CSF for neutropenia.

This study is not without its limitations. The study ultimately included a modest number of patients receiving G-CSF (N=16). In this observational cohort, unaccounted confounding factors are plausible. This analysis attempted to assess G-CSF over a wide range of cancers; tumor-specific effects were difficult to ascertain. We also limit our primary analysis to the subset of patients who were hospitalized, to be able to assess our clinical endpoints of interest (respiratory failure or death). As a result, the potential for G-CSF to worsen outcomes, suggested here, may not be generalizable to outpatients. While our analysis adjusts for the neutrophil count prior to G-CSF administration, we have not incorporated data on concurrent therapies such as chemotherapies or surgery prior to the diagnosis of COVID-19 in these patients.

Interestingly, in lieu of the COVID-19 pandemic, the National Comprehensive Cancer Network and the American Society of Clinical Oncology have released updated guidelines on this very issue, lowering the threshold for the use of G-CSF to now include those chemotherapy regimens that carry a 10 to 20 percent risk of fever in the setting of neutropenia^24^. Although there is a potential role for G-CSF and its prophylactic use following the administration of chemotherapy when neutropenia is anticipated (primary prophylaxis) and during retreatment after a previous cycle of chemotherapy had caused fever during neutropenia (secondary prophylaxis),^25^ G-CSF has not definitively been shown to actually reduce infection-related mortality. ^26,27,28^ Its benefits therefore must be carefully weighed against the potential risk of harm in the patient with cancer and active COVID-19 infection, particularly one requiring hospitalization. Until additional data from larger studies are available, in areas with high or increasing incidence of COVID-19, perhaps the use of G-CSF in patients with confirmed COVID-19 could be reserved for those patients who are classified as having a higher risk of complications- those that have profound neutropenia due to chemotherapy. ^29^

## Data Availability

The data that support the findings of this study are available on request from the corresponding author, [SM]. The data are not publicly available due to [restrictions e.g. their containing information that could compromise the privacy of research participants].

## Acknowledgements

The authors would like to thank Susan Weill who works in the Design and Creative Services, Marketing and Communication Department at MSKCC who helped to create the figures for this manuscript.

---

**Figure S1:**
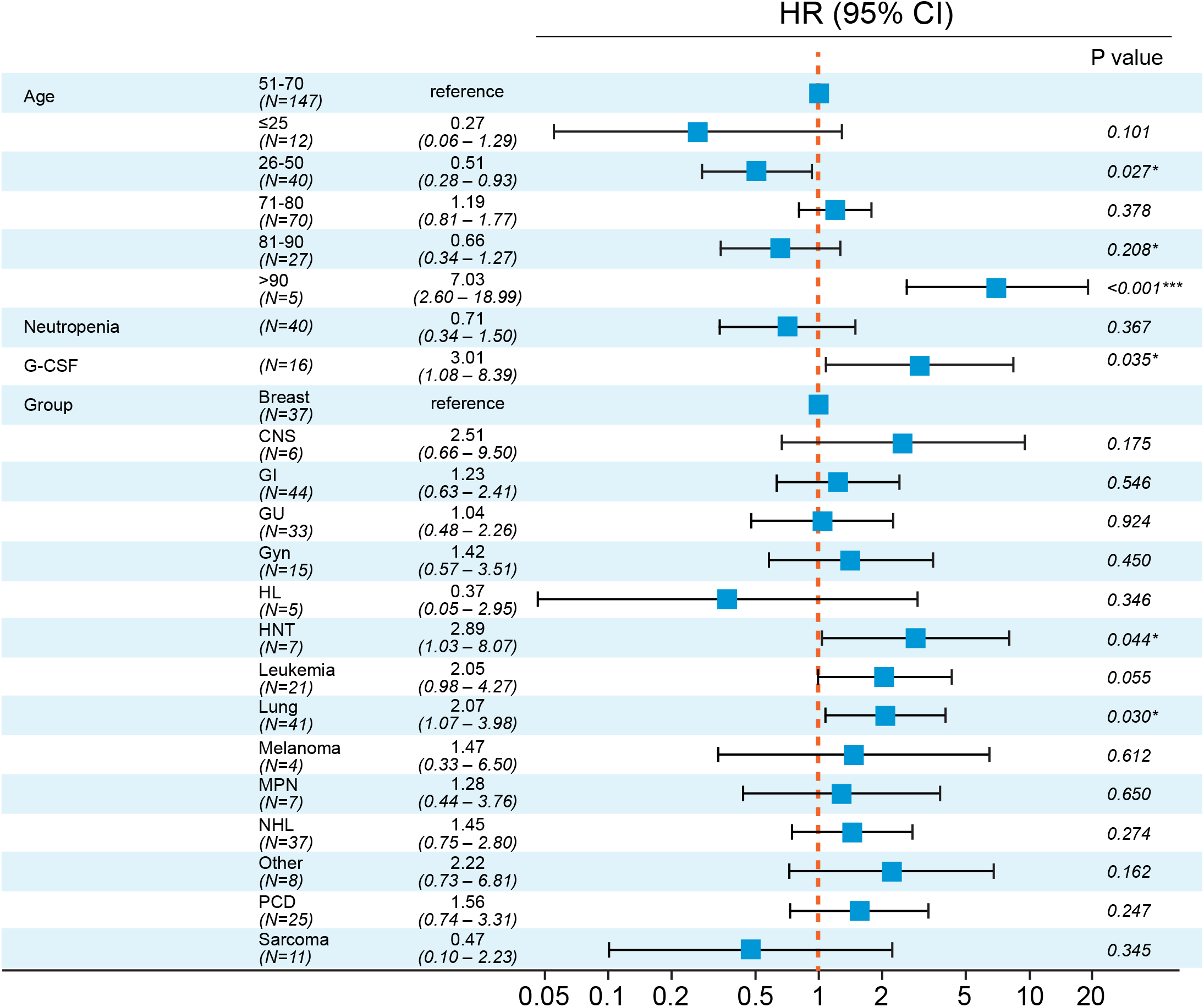
Forest plot showing the effect (HR = Hazard Ratio) of Granulocyte colony-stimulating factor (G-CSF) on the first occurrence of “respiratory failure” (defined in Methods). Hazard ratios were computed with an extended Cox model, using binned ages and cancer type as time-independent covariates and neutropenia and G-CSF as time-dependent covariates.

**Figure S2:**
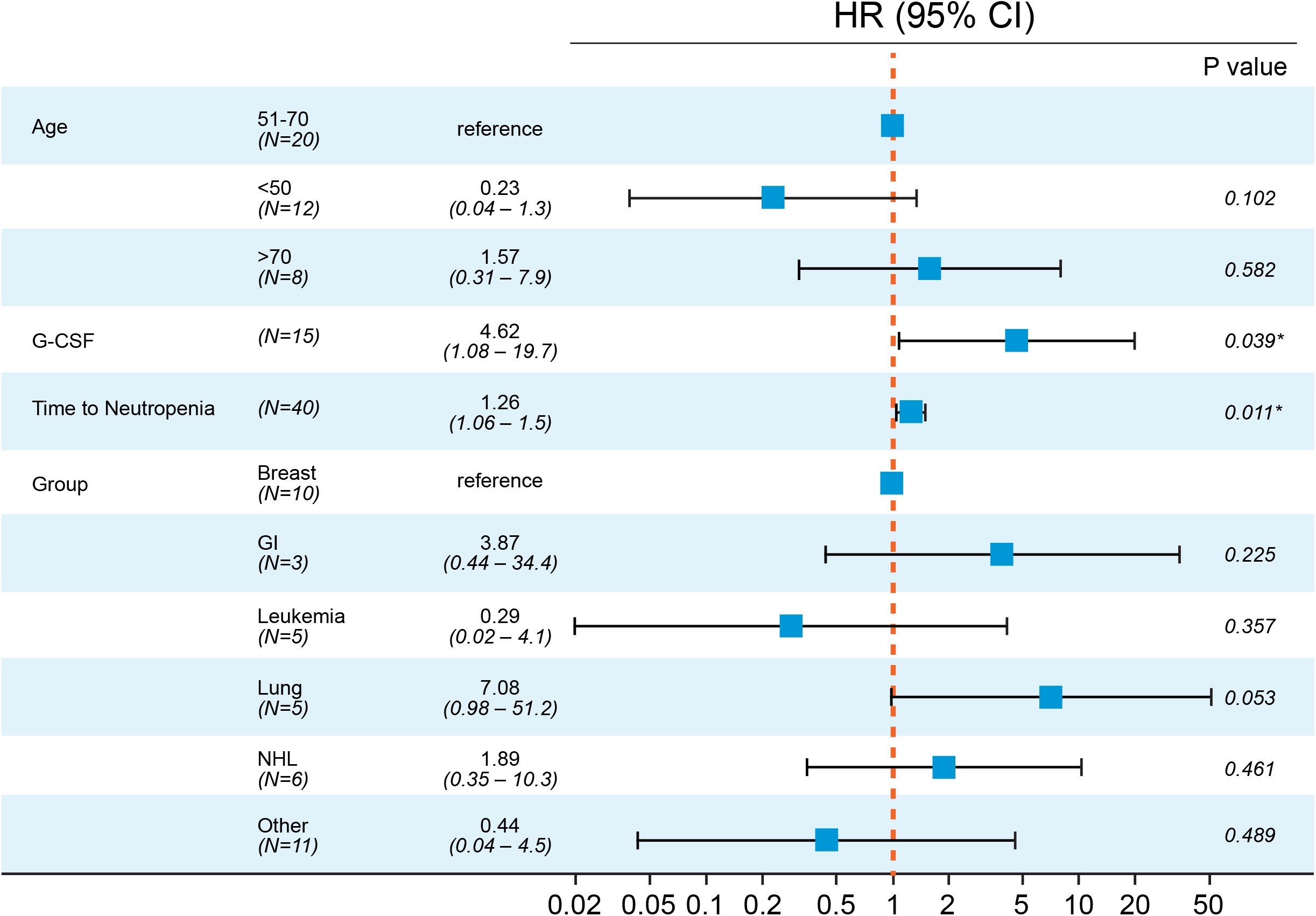
Forest plot showing the effect (HR = Hazard Ratio) of G-CSF administration on the first occurrence of “respiratory failure” (defined in Methods), using the time of the first occurrence of neutropenia as t=0. Hazard ratios were computed with an extended Cox model, using binned ages, cancer type, and the time to neutropenia (from COVID-19+ date) as time-independent covariates and G-CSF as a time-dependent covariate.

**Figure S3:**
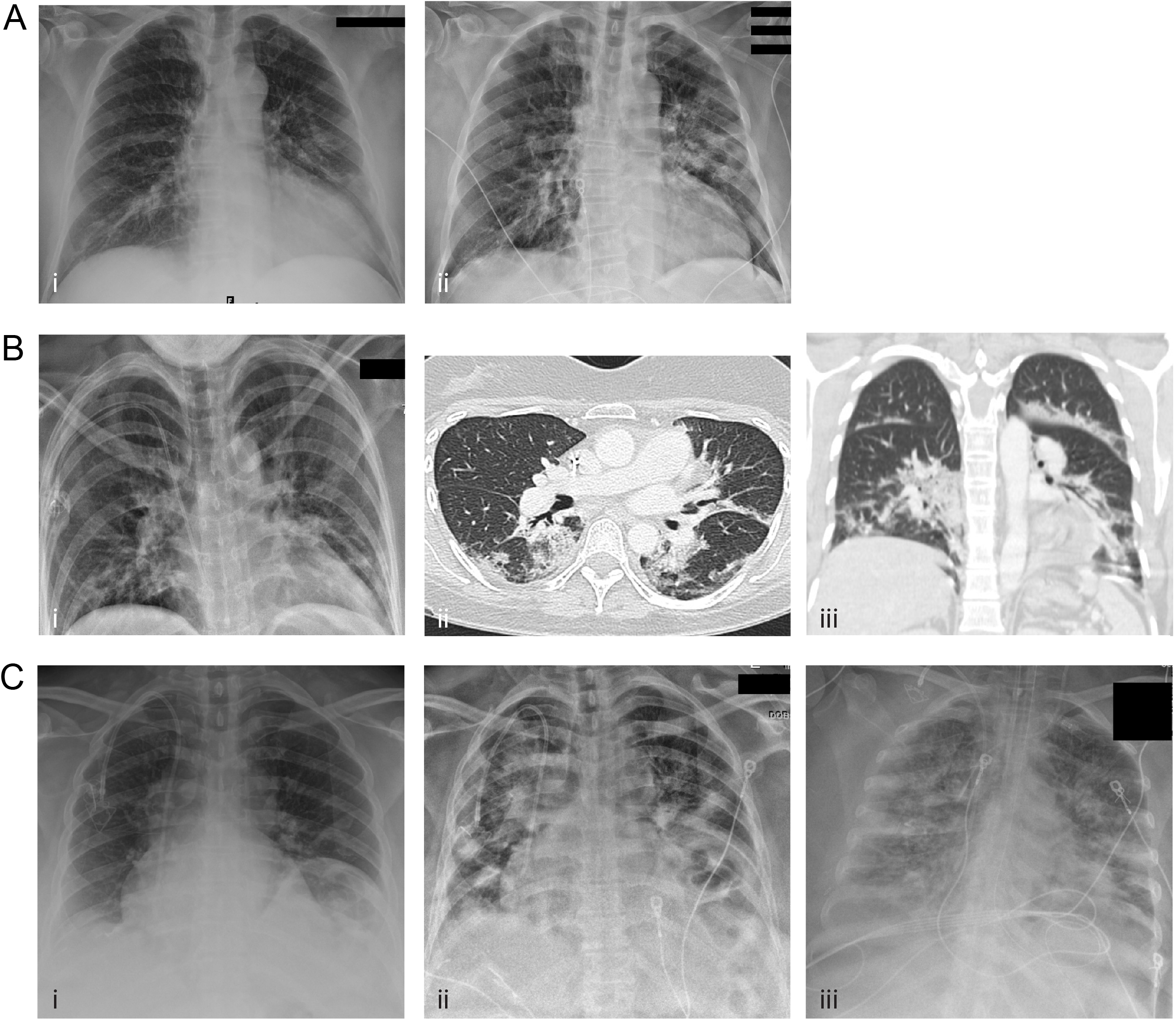
**A** i: Portable chest X-Ray performed the day of G-CSF administration demonstrating right basilar and left mid lung patch opacities. A ii: A day after the administration the airspace opacities increased bilaterally **B** i: Portable chest X-Ray performed two days prior to administration of G-CSF demonstrating bilateral predominantly bibasilar patchy opacities B ii and B iii: Axial and coronal images two days post G-CSF administration, demonstrating peripheral and peribronchovascular airspace opacities predominantly in the lower lobes **C** i: Portable chest X-ray at day 0 of G-CSF administration demonstrating faint right basilar opacity. C ii: day 4 post G-CSF administration, bilateral patchy opacities noted involving both upper and lower zones C iii: Day 13 post administration, increased bilateral diffuse reticular and airspace opacities

**Table S1:** Demographics and baseline characteristics of neutropenic patients that did (n= 29) and did not receive G-CSF (n= 55).

| Characteristic | Neutropenia (N=55), n (%) | G-CSF use (N=29), n (%) |
| --- | --- | --- |
| **Age (years)** |  |  |
| < 18 years | 7 (12.7) | 5 (17.2) |
| 18-29 | 1 (1.8) | 0 |
| 30-39 | 1 (1.8) | 1 (3.5) |
| 40-49 | 6 (10.9) | 4 (13.8) |
| 50-59 | 17 (30.9) | 8 (27.6) |
| 60-69 | 13 (23.6) | 6 (20.7) |
| >70 years | 10 (18.2) | 5 (17.2) |
| **Gender** |  |  |
| Male | 36 (65.5) | 24 (82.8) |
| Female | 19 (35.5) | 5 (17.2) |
| **Race** |  |  |
| White | 31 (56.4) | 14 (48.3) |
| Black | 12 (21.8) | 8 (27.6) |
| Asian | 2 (3.6) | 1 (3.5) |
| Other/Inknown | 10 (18.2) | 6 (20.7) |
| **Underlying Cancer\*** |  |  |
| Breast cancer | 10 (18.2) | 8 (27.6) |
| CNS<sup>+</sup> | 1 (1.8) | 1 (3.5) |
| Lung cancer | 6 (10.9) | 3 (10.3) |
| Gastrointestinal malignancies<sup>∞</sup> | 4 (7.3) | 2 (6.9) |
| Genitourinary malignancies<sup>Δ</sup> | 2 (3.6) | 0 |
| Gynecologic malignancies<sup>∇</sup> | 2 (3.6) | 1 (3.5) |
| Melanoma | 1 (1.8) | 1 (3.5) |
| Sarcoma | 2 (3.6) | 2 (6.9) |
| Acute and chronic leukemias<sup>†</sup> | 7 (12.7) | 3 (10.3) |
| Plasma cell dyscrasias<sup>‡</sup> | 6 (10.9) | 3 (10.35) |
| Myeloproliferative neoplasms<sup>▷</sup> | 1 (1.8) | 0 |
| Non-Hodgkin lymphoma | 12 (21.8) | 5 (17.2) |
| Other | 1 (1.8) | 0 |

**CNS cancers<sup>+</sup>** = Astrocytoma, glioblastoma, glioma, nerve sheath tumor, neuroblastoma
**Germ cell tumors<sup>Δ</sup>** = Germ cell tumor and testicular cancer
**Gastrointestinal malignancies<sup>∞</sup>** = Appendiceal, cholangiocarcinoma, gallbladder, gastrim, esophageal, hepatocellular, pancreatic, rectal/colon
**Genitourinary malignancies<sup>Δ</sup>** = Bladder cancer, urothelial cancer, prostate cancer, renal cell carcinoma
**Gynecologic malignancies<sup>∇</sup>** = cervical, endometrial, ovarian, uterine, vulvar
**Acute and chronic leukemias<sup>†</sup>** = acute lymphoblastic leukemia, acute myeloid leukemia, chronic lymphoid leukemia, chronic myeloid leukemia, chronic myelomonocytic leukemia, hairy cell leukemia, myelodysplastic syndrome
**Plasma cell dyscrasias<sup>‡</sup>** = amyloidosis, multiple myeloma, monoclonal gammopathy of undetermined significance

**Table S2:** Demographics and baseline characteristics of all outpatients with COVID-19 (n = 530)

| Characteristic | Number of Patients, n (%) |
| --- | --- |
| **Age (years)** |  |
| < 18 years | 12 (2.26) |
| 18-29 | 23 (4.34) |
| 30-39 | 39 (7.36) |
| 40-49 | 76 (14.3) |
| 50-59 | 133 (25.1) |
| 60-69 | 140 (26.4) |
| > 70 | 107 (20.2) |
| **Gender** |  |
| Male | 263 (49.6) |
| Female | 267 (50.4) |
| **Race** |  |
| White | 325 (61.3) |
| Black | 89 (16.8) |
| Asian | 51 (9.62) |
| Other/Unknown | 64 (12.1) |
| **Underlying Cancer\*** |  |
| Breast cancer | 105 (19.8) |
| Nervous system cancers | 10 (1.89) |
| Germ cell tumors | 8 (1.51) |
| Lung cancer | 33 (6.23) |
| Gastrointestinal malignancies | 90 (17.0) |
| Genitourinary malignancies | 71 (13.4) |
| Gynecologic malignancies | 24 (4.52) |
| Head and neck cancers | 23 (4.33) |
| Skin cancer | 15 (2.83) |
| Sarcoma | 35 (6.6) |
| Acute and chronic leukemias | 47 (8.86) |
| Plasma cell dyscrasias | 22 (4.15) |
| Myeloproliferative neoplasms | 4 (0.75) |
| Non-Hodgkin's and Hodgkin's lymphoma | 41 (7.73) |
| Other | 2 (0.4) |
| **Outpatient G-CSF use** |  |
| Filgrastim | 11 (1.7) |
| Pegylated Filgrastim | 1 (0.2) |
| Neither | 518 (97.7) |
| **Death** | 21 (3.4%) |

\***Nervous system cancers** include glioma, glioblastoma, astrocytoma, nerve sheath tumors, neuroblastoma; germ cell tumors include germ cell tumors and testicular cancer; **gastrointestinal malignancies** include oropharyngeal cancer, appendiceal cancer, cholangiocarcinoma, gallbladder cancer, gastric cancer, esophageal cancer, hepatocellular carcinoma, pancreatic cancer, colorectal cancer, gastrointestinal stromal tumors, pancreatic neuroendocrine tumors; **genitourinary malignancies** include bladder cancer, urothelial cancer, prostate cancer, renal cell carcinoma; gynecologic malignancies include cervical cancer, ovarian cancer, endometrial cancer, uterine cancer, vulvar cancer; **head and neck cancers** include head and neck squamous cell carcinoma, adenoid carcinoma, thyroid cancer, thymic tumors; **skin cancers** include melanoma and non-melanoma skin cancer; **acute and chronic leukemias** include acute and chronic myeloid leukemia, acute and chronic lymphocytic leukemia, hairy cell leukemia, myelodysplastic syndrome, chronic myelomonocytic leukemia; **plasma cell dyscrasias** include multiple myeloma, amyloidosis, smoldering myeloma, monoclonal gammopathy of undetermined significance; **myeloproliferative neoplasms** include myelofibrosis, immune thrombocytopenic purpura, aplastic anemia, Erdheim-Chester syndrome.

**Table S3:** Descriptive statistics (frequencies as %) of both inpatients and outpatients that did and did not receive G-CSF and non-G-CSF with clinical endpoint. This does not account for time dependence of G-CSF risk as primary analysis does

|  | No G-CSF | G-CSF | G-CSF use prior to combined endpoint only |
| --- | --- | --- | --- |
| Outpatient |  |  |  |
| Death | 21/518 = 4% | 0/12 = 0% | N/A |
| Inpatient |  |  |  |
| High O2 (>4L) | 94/275 = 34% | 9/29 = 31% | 7/16 = 44% |
| Vent | 47/275 = 17% | 4/29 = 14% | 1/16 = 6% |
| Death | 49/275 = 18% | 4/29 = 14% | 2/16 = 13% |

**Table S4:** Radiologic evolution of patients receiving G-CSF. Baseline X-Ray of patients was determined as normal or abnormal if airspace or reticulonodular opacities were noted. X-Ray post G-CSF was compared to baseline assessing radiologic evolution and categorizing it as: unchanged, increased or decreased.

| Patient | Baseline Chest Xray:<br>Normal (0) vs<br>Abnormal (1)<br>Pre- G-CSF | Follow up<br>Chest Xray:<br>Yes vs. No<br>Post- G-CSF | Days between<br>G-CSF and<br>follow up<br>Chest Xray | Evolution |
| --- | --- | --- | --- | --- |
| 1 | 1 | Yes | 2 | Increased |
| 2 | 0 | No | NA | NA |
| 3 | 1 | Yes | 13 | Decreased |
| 4 | 0 | Yes | 0 | Increased |
| 5 | 1 | Yes | 2 | Increased |
| 6 | NA | No | NA | NA |
| 7 | 1 | Yes | 2 | Decreased |
| 8 | 0 | Yes | 2 | Increased |
| 9 | 1 | Yes | 2 | Decreased |
| 10 | 0 | Yes | 0 | Unchanged |
| 11 | 1 | Yes | 5 | Increased |
| 12 | 0 | Yes | 1 | Increased |
| 13 | 1 | NA | NA | NA |
| 14 | 1 | NA | NA | NA |
| 15 | 1 | NA | NA | NA |
| 16 | NA | NA | NA | NA |

